# Ancestry-specific immune signatures in Parkinson’s disease: a rare variant burden analysis in Montreal and Guadeloupe cohorts

**DOI:** 10.64898/2026.04.24.26351405

**Authors:** Lovatiana Andriamboavonjy, Marjorie Labrecque, Lalla Yasmine Al Idrissi, Benoît Tressières, Ariane Veilleux Carpentier, Juliette Chaumont, Hugo Chaumont, Stanie Gaete, Sylvie Ravion, Antoine Duquette, Sylvain Chouinard, Michel Panisset, Annie Lannuzel, Martine Tetreault

**Affiliations:** CHUM Research center, Montreal, QC, Canada; Department of Neurosciences, Université de Montréal, Montreal, QC, Canada; Department of Medicine, Université de Montréal, Montreal, QC, Canada; Caribbean Clinical Investigation Center, Inserm CIC 2504, University hospital of Guadeloupe, France; Department of Neurology, University hospital of Guadeloupe, France; Karubiotec™, Biological Resources Center, University Hospital of Guadeloupe, France; Unité des troubles du mouvement André-Barbeau, Centre Hospitalier de l’Université de Montréal, Montreal, Canada; Department of Medicine Antilles-Guyane, Antilles University, France; Paris Brain Institute-ICM, Inserm, CNRS, Hôpital de la Pitié Salpêtrière, Sorbonne Université, 75013 Paris, France

**Keywords:** Parkinson’s disease, RNA-sequencing, Rare variant burden, Ancestry-specific mechanisms, Immunity, Linkage disequilibrium

## Abstract

**Background:** Parkinson’s disease (PD) genetics research has predominantly focused on populations of European ancestry, limiting understanding of disease mechanisms across diverse populations. African-Caribbean communities harbor complex genetic admixture and exhibit distinct clinical features, yet remain underrepresented in PD genetic studies. We investigated ancestry-specific molecular signatures underlying PD using rare variant burden analysis in geographically and genetically distinct cohorts.

**Methods:** Using a discovery cohort design and gene-based rare-variant aggregation testing, we performed RNA-sequencing on peripheral blood mononuclear cells from 33 participants: Montreal, Canada (n=16; 8 PD, 8 controls) and Guadeloupe, French West Indies (n=17; 9 PD, 8 controls). We conducted gene-based rare variant burden testing, protein-protein interaction network analysis, pathway enrichment, and linkage disequilibrium (LD) profiling. Clinical assessments included MDS-UPDRS, Hoehn & Yahr staging, and evaluation of prodromal and autonomic features.

**Results:** Principal component analysis revealed distinct population structure, with Montreal participants forming a homogeneous cluster and Guadeloupe participants displaying greater variance consistent with African-European admixture. Ancestry-stratified burden analysis identified divergent immune pathway enrichments: IL-17 signaling predominated in PD patients from Montreal (FDR=0.04), while MHC class II antigen presentation and interferon-γ pathways characterized PD patients from Guadeloupe (FDR=1.59×10⁻⁷). A *NOD2* frameshift variant (rs2066847) was identified in 3/8 Montreal patients, providing a mechanistic link to IL-17 pathway dysregulation. LD analysis revealed ancestry-specific haplotype structures, with 14 African-admixed American-specific LD pairs exclusive to Guadeloupe participants and 11 European-specific pairs present in both populations, demonstrating distinct haplotype architectures shaped by ancestry. Clinical differences aligned with molecular findings: Montreal patients showed higher prevalence of REM sleep behavior disorder (71.4% vs 37.5%) and hyposmia (54.3% vs 22.5%), while Guadeloupe patients showed more autonomic symptoms. Control-only comparison showed no pathway enrichments, validating that PD findings reflect disease-associated mechanisms rather than population stratification.

**Conclusions:** Together, these findings provide evidence for ancestry-specific immune signatures in PD, challenging a one-size-fits-all paradigm in neurodegenerative disease genetics. The identification of distinct molecular pathways underlying clinically overlapping phenotypes suggests PD may encompass multiple molecular entities converging on shared symptoms. These findings emphasize the necessity of ancestry-inclusive research for advancing mechanistic understanding and achieving equity in precision medicine for neurodegenerative disorders.

## BACKGROUND

Parkinson’s disease (PD) genetics research has mostly focused on populations of European ancestry over the past decade [1,2,3]. This Eurocentric bias limits our understanding of disease mechanisms across diverse populations, as genetic risk variants and their functional consequences may differ substantially across ancestries [4,5]. The resulting knowledge gaps have profound implications for precision medicine, as molecular mechanisms identified in one population may not generalize to others [6].

Among underrepresented populations, African-Caribbean communities have complex genetic admixture resulting from historical migration and colonization, with substantial contributions from West African, European, and Indigenous American ancestries [7,8]. Interestingly, a form of atypical parkinsonism characterized by bradykinesia-dominant presentation, reduced response to levodopa, and earlier cognitive decline has been specifically described in the French Antilles, highlighting potential population-specific variation in motor and non-motor symptoms of degenerative parkinsonism [9,10].

While common variants identified through Genome-wide association study (GWAS) explain only part of PD heritability [11], low-frequency coding variants with larger effect sizes may contribute substantially to disease risk [12]. Furthermore, canonical monogenic PD genes such as *LRRK2* account for only a small proportion of sporadic PD cases [13], suggesting that alternative genetic mechanisms may contribute to idiopathic PD pathogenesis.

Neuroinflammation, including microglial activation and astrogliosis, is well documented in PD [14,15,16]. In addition, recent studies suggest potential involvement of the peripheral immune system alterations, with observations of altered T cell, B cell, and monocyte profiles in some PD patient cohorts [17,18,19]. Whether these peripheral immune changes represent causal mechanisms, consequences of disease, or population-specific events remains unclear and emphasizes the need for systematic investigation across ancestries.

To address these gaps, we used a discovery cohort design and performed gene-based variant burden testing. Because rare coding variants with moderate effect sizes may contribute to disease risk, gene-based aggregation approaches provide greater statistical power than single-variant tests. This analysis was followed by protein-protein interaction (PPI) network construction, pathway enrichment analysis, and linkage disequilibrium (LD) profiling in idiopathic PD patients and healthy controls (CTRL) from two geographically distinct populations: Montreal, Canada and Guadeloupe, French West Indies. These populations are expected to differ in genetic ancestry, with Montreal participants predominantly of European descent and Guadeloupe participants representing admixed African-Caribbean ancestry. Using peripheral blood mononuclear cell (PBMC) RNA sequencing, we aimed to compare genetic architecture within and across these populations to identify ancestry-specific molecular mechanisms underlying PD.

## METHODS

The study workflow is summarized in **Figure 1**.

**Figure 1.**
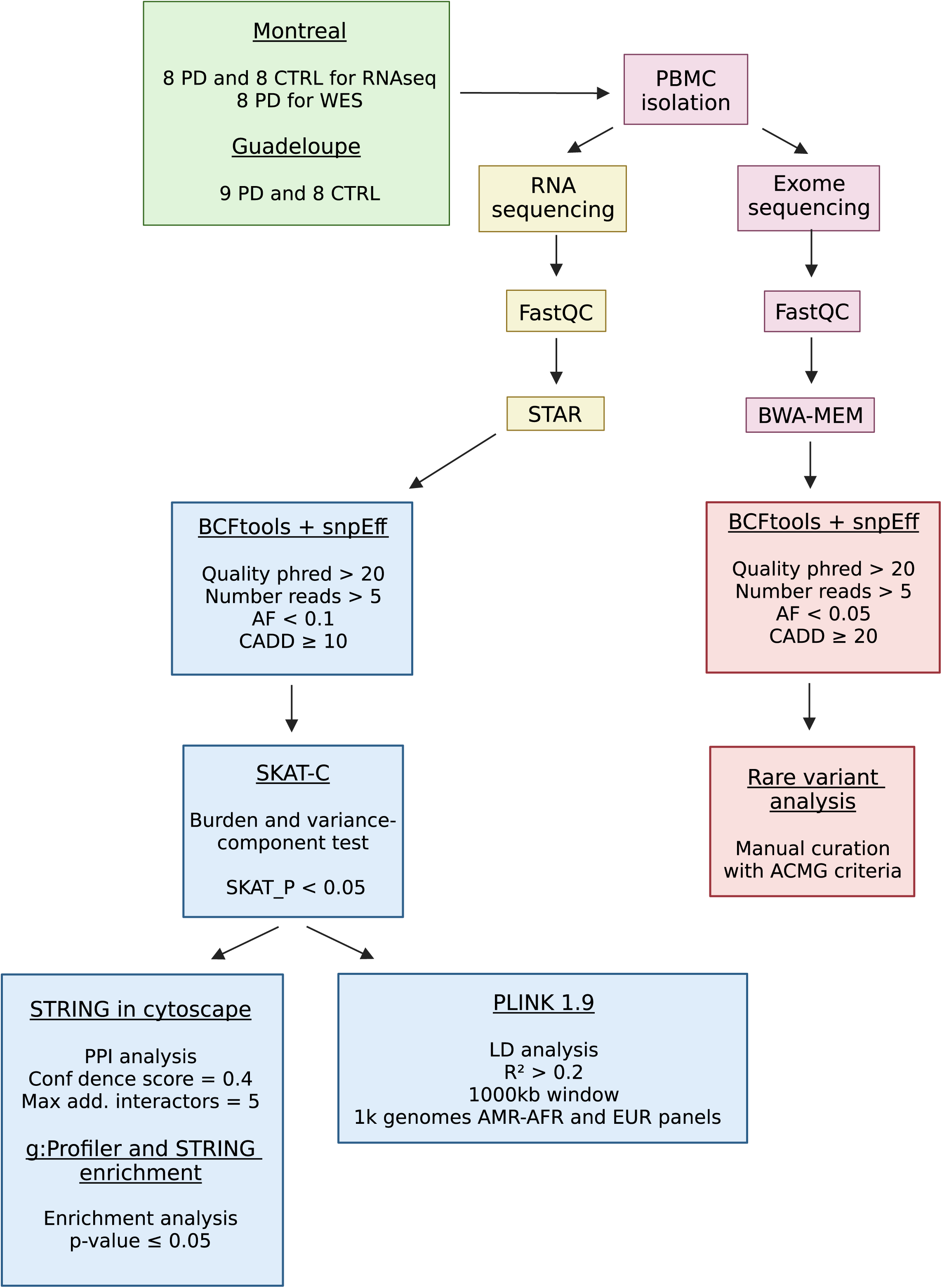
Analytical workflow for ancestry-specific rare variant analysis in Parkinson’s disease (PD). Schematic overview of the study design and bioinformatics pipeline. Peripheral blood mononuclear cells (PBMC) were isolated from participants in Montreal (8 PD and 8 controls for RNA-seq; 8 PD for whole exome sequencing) and Guadeloupe (9 PD and 8 controls for RNA-seq). RNA-seq data were processed using FastQC for quality control and STAR for alignment, followed by variant calling with BCFtools and FreeBayes and annotation with snpEff. Variants were filtered (quality phred > 20, read depth > 5, allele frequency < 0.1, CADD ≥ 10) and analyzed using SKAT-C for burden and variance-component testing (p < 0.05). Significant genes underwent protein-protein interaction (PPI) network analysis in STRING/Cytoscape (confidence score = 0.4, maximum 5 additional interactors) and pathway enrichment analysis using g:Profiler and STRING (p ≤ 0.05). Linkage disequilibrium (LD) analysis was performed using PLINK 1.9 (R² > 0.2, 1000 kb window) with 1000 Genomes Project AFR-AMR and EUR reference panels. Whole exome sequencing data were processed using BWA-MEM for alignment with more stringent filtering criteria (AF < 0.05, CADD ≥ 20) for rare variant analysis and manual curation following ACMG criteria.

### Study participants and sample collection

This discovery cohort included 33 participants recruited from two sites: Montreal, Quebec, Canada (abbreviated **MTL**; n=16; 8 PD, 8 CTRL, age- and sex-matched) and Pointe-à-Pitre, Guadeloupe, French Antilles (abbreviated **PTP**; n=17; 9 PD, 8 CTRL, age- and sex-matched). Recruitment spanned 2020 to 2023. All patients met diagnostic criteria based on the Movement Disorders Society (MDS) and current consensus [20]. CTRL were individuals without neurological disorders or a family history of PD. Patients with clinical features suggestive of atypical parkinsonism were excluded from the study.

Blood samples were collected after informed written consent was obtained in accordance with the institutional Ethics Committee Protocol #20.367 and #2020-88

### PBMC isolation

Blood was collected in ethylenediaminetetraacetic acid (EDTA) Vacutainer tubes and processed at room temperature within 4 hours of collection. Whole blood was diluted with phosphate-buffered saline (PBS)-2 mM EDTA in 50 mL tubes and layered over 12 mL of Ficoll-Paque (GE Healthcare). Following density gradient centrifugation according to the manufacturer’s instructions, the buffy coat containing PBMCs was harvested and washed with PBS-2 mM EDTA. PBMCs were aliquoted for long-term storage in liquid nitrogen in a freezing solution containing 10% dimethyl sulfoxide (Sigma-Aldrich) in fetal bovine serum (Gibco).

### RNA extraction and quality control

Total RNA was extracted from PBMCs using the RNAeasy mini kit 250 prep from Qiagen (Catalog 74104) according to the manufacturer’s protocol. RNA quality and integrity were assessed using the TapeStation system (Agilent Technologies). Due to variability in PBMC processing and storage, we included samples with RNA Integrity Number (RIN) ≥ 6, a threshold validated for biobanked brain tissue [21]. Although this threshold is lower than conventional thresholds for high-quality RNAseq, prior studies suggest that mRNA integrity can remain sufficient for transcriptomic analyses despite partial degradation [22]. To confirm this threshold did not introduce technical bias, we verified that RIN values showed weak correlation (R² = 0.09; P = 0.04) with the number of mapped reads in our dataset **(Supplementary figure 1)** and observed no systematic differences in RIN distribution between the MTL and PTP cohorts.

### RNA Sequencing

RNA sequencing libraries were prepared using the NEBNext Ultra II Directional RNA library Prep Kit with NEBNext Poly(A) mRNA-dual index following the manufacturer’s instructions. Libraries were sequenced on an Illumina NovaSeq 6000 S4 flowcell to generate approximately 50 million 100 base pair paired-end reads per sample. Sequencing was performed at the Centre Génomique du CHU de Quebec - Université Laval.

### Bioinformatics Pipeline

#### Read Processing and Alignment

Raw sequencing reads were assessed for quality using FastQC version 0.12.1 [23]. High-quality reads were aligned to the human reference genome (GRCh38) using the STAR aligner version 2.7.11 [24].

#### Variant Calling and Annotation

Variant calling from RNA-seq alignments was performed using BCFtools version 1.18 [25] and FreeBayes version 1.37 [26]. Variants were functionally annotated using snpEff version 5.2 [27,28], which provides consequence predictions, amino acid changes, and allele frequency information from integrated population databases. This study does not aim to comprehensively characterize all genetic variation, but to interrogate expressed and functionally relevant variation in immune cells. RNA-seq-derived variants therefore reflect transcriptionally active loci, enriching for variation with direct functional consequences in the peripheral immune compartment relevant to our research question.

#### Variant Quality Control and Filtering

Stringent quality control filters were applied for appropriate burden testing: base quality (phred score) ≥ 20; minimum read depth ≥ 20 supporting reads; alternate allele frequency (AF) ≤ 10% in reference populations (gnomAD) to capture rare and low-frequency variants. To enrich for variants with potential functional impact, we retained variants with a CADD phred score ≥ 10, indicating variants predicted to be among the 10% most deleterious. Because variant calling was performed on RNA-seq alignments, analyses were intentionally restricted to well-supported variants in expressed transcripts, prioritizing functional relevance over exhaustive variant discovery. The inclusion of low-frequency variants (AF 1-10%) alongside rare variants is explained by the complex polygenic nature of PD. Multiple variants of modest individual effect may collectively contribute to disease risk and phenotypic heterogeneity.

#### Principal component analysis

Population structure was assessed using principal component analysis (PCA) of genetic variants in R version 4.5.2. Samples were annotated by disease phenotype (PD vs CTRL) and ancestry (MTL/European vs PTP/African-Caribbean) to visualize population stratification. Mean pairwise genetic distances were calculated as the mean absolute difference in genotype calls across all variants between each pair of samples to quantify genetic diversity within and between populations.

#### Genotype matrix construction

For each comparison, a genotype matrix was constructed from the filtered variants. Samples were grouped based on disease status (PD vs CTRL) and site of origin (MTL, PTP). The genotype matrix was generated with values representing genotype calls (0 = reference, 1 = heterozygous, 2= homozygous alternate). Missing samples (those with no variants passing filters) were inserted as all-zero columns to maintain the complete sample set. A variant-to-gene mapping file was generated concurrently, associating each variant identifier with its corresponding gene name.

#### SKAT-C rare variant burden testing

Given the modest cohort size, we applied SKAT-C for gene-based aggregation of rare and low-frequency variants, a method specifically designed for burden testing and well suited for small case-control datasets.

Three primary comparisons were performed: **(1)** PD_MTL vs CTRL_MTL, **(2)** PD_PTP vs CTRL_PTP, **(3)** PD_MTL vs PD_PTP. We also performed a CTRL_MTL vs CTRL_PTP analysis to differentiate ancestry-related and disease-related results. Gene-level rare variant burden testing was performed using Sequence Kernel Association Test-CommonRare (SKAT-C) from the SKAT R package v.2.2.5 [29]. A null model was constructed using a binary phenotype (case = 1, control = 0). For each gene with at least two variants, SKAT-C analysis was performed using the “SKAT_CommonRare()” function with the following parameters: rare variant weights beta = c(1,25), common variant weights beta = c(0.5, 0.5), common/rare AF cutoff = 0.01, and test type = “Joint”. No covariates were included in the SKAT-C analysis because PD patients and CTRL from both MTL and PTP cohorts were already matched for age and sex **(Table 1)**. To preserve potential ancestry-specific signals, we did not adjust the analysis for population structure.

**Table 1.** Participant demographics for RNAseq and whole-exome sequencing cohorts. Age is reported as mean ± standard deviation. PD: Parkinson’s disease; CTRL: Control; PTP: Pointe-à-Pitre; MTL: Montreal.

| | n | Female | Male | Age, mean ± SD (years) |
| --- | --- | --- | --- | --- |
| **RNA sequencing** |  |  |  |  |
| *PTP cohort* |  |  |  |  |
| PD | 9 | 6 | 3 | 63.1 ± 5.8 |
| CTRL | 8 | 5 | 3 | 63.1 ± 6.0 |
| *MTL cohort* |  |  |  |  |
| PD | 8 | 6 | 2 | 65.0 ± 4.0 |
| CTRL | 8 | 5 | 3 | 64.5 ± 3.9 |
| **Whole-exome sequencing** |  |  |  |  |
| *MTL cohort* |  |  |  |  |
| PD | 8 | 1 | 7 | 64.0 ± 9.4 |

For each gene, the following metrics were calculated: SKAT-C p-value, statistical significance of the gene-level burden; Number of rare variants, variants with MAF < 0.01; Number of common variants, variants with MAF ≥ 0.01; Delta burden, difference in mean variant count between cases and controls; directional score, sign(Δburden) × -log₁₀(p-value), indicating the comparison group variant-enriched. Genes were ranked by SKAT-C p-value, and a significance threshold of nominal P ≤ 0.05 was applied to identify genes with significant mutation burden differences between groups.

#### Protein-protein interaction network construction

PPI networks were constructed for significantly burdened genes, using stringApp v2.2.0 integrated in Cytoscape v3.10.3. Network parameters included: minimum required interaction confidence score = 0.4, minimum degree = 2, and five additional interactors, focusing on high-confidence direct interactions. **(Figure 1)**

#### Pathway enrichment analysis

Significant genes identified in SKAT-C analysis were subjected to functional enrichment analysis using g:Profiler (accessed through EnrichmentTable v2.0.7 in Cytoscape) and STRING enrichment analysis version 12.0 (through stringApp v3.1.0). g:Profiler tested for over-representation in KEGG, Reactome, and Gene Ontology (GO) pathways, while STRING enrichment tested STRING clusters, Wikipathways, UniProt keywords and Diseases. Statistical significance was defined as FDR ≤ 0.05, controlling for multiple testing across all tested pathways **(Figure 1)**. To reduce redundancy in pathway enrichment results, pathways were merged using Jaccard similarity clustering. For each enrichment source, pathways were ranked by false discovery rate (FDR). Starting from the most significant pathway, Jaccard similarity coefficients were computed pairwise between gene sets (J = |A ∩ B| / |A ∪ B|). Pathways with Jaccard similarity > 0.7 were clustered together. For each cluster, the representative pathway was selected prioritizing database hierarchy and lowest FDR. The merged pathway label reflected the representative term. Statistical significance for merged clusters was defined as the minimum FDR across all merged terms.

#### Linkage Disequilibrium Analysis

To identify ancestry-specific haplotype structures potentially contributing to phenotypic differences between populations, we performed LD analysis between variants from SKAT-C-significant genes identified in our cohorts and variants from the 1000 Genomes Project Phase 3 reference panels (release 20130502; 1000 Genomes Project Consortium, 2015).

##### LD Calculation

To identify ancestry-specific haplotype structures, we first performed SKAT-C analysis combining all participants (PD_ALL vs CTRL_ALL, n=33) to identify genes with significant variant burden regardless of ancestry. Variants from these significant genes were then used as query variants for LD analysis. Pairwise LD (R²) was calculated using PLINK v1.9 within a 1000 kb sliding window around each query variant, computed separately against two 1000 Genomes Project Phase 3 reference panels: European ancestry (EUR) and combined African-admixed American (AFR-AMR).

##### LD filtering and annotation

Raw LD results were filtered to retain only variant pairs with R^2^≥ 0.8, excluding self-pairs. Study variants (SNP_A) were required to be present in the genotype matrix, while linked variants (SNP_B) were required to be from the 1000 Genomes reference panel. Bidirectional duplicate pairs were retained to preserve information about haplotype structure.

Variants were annotated with gene names, using the study genotype matrix. If the variant was missing in the matrix, unmapped variants were annotated using GENCODE v48 annotation (GRCh38).

#### Clinical assessment

PD patients in this genomics study represent a subset of a larger clinically characterized cohort including 135 patients: 66 from MTL and 69 from PTP **(Supplementary Table 1)**. All patients underwent comprehensive clinical evaluation by movement disorder specialists at their respective sites. Clinical assessments were standardized across both sites and included the Movement Disorder Society-Unified ’s Disease Rating Scale (MDS-UPDRS) [28] to evaluate motor symptoms, non-motor experiences of daily living, and motor complications. Disease stage was determined using the Hoehn and Yahr scale. Additional assessments included evaluation of prodromal features such as REM sleep behavior disorder (RBD), assessed using a validated single-question screen [30], and self-reported hyposmia, as well as autonomic symptoms including orthostatic hypotension. Clinical data were collected at the time of blood sampling for RNA sequencing to ensure temporal alignment between molecular and phenotypic assessments.

#### Whole-exome sequencing analysis

We isolated PBMC from eight PD patients recruited from an independent cohort **(Table 1)**, distinct from the RNAseq cohort, at the Movement Disorder Unit at CHUM, Montreal, and diagnosed according to MDS-UPDRS criteria, using the same PBMC isolation protocol as the RNAseq analysis. Genomic DNA was extracted using the RNA/DNA/Protein purification plus kit from Norgen Biotek (catalog 47700) and whole-exome sequencing was performed on a NovaSeq 6000 platform (25M PE100 reads per sample). Quality control of raw reads was performed using FastQC version 0.12.1 [23]. Reads were aligned to a reference genome (GRCh38) using BWA-MEM version 2.2.1 [31] while variant calling was done the same way as the RNA-seq. For rare variant analysis, we applied the following filters: base quality phred score > 20, read depth (> 5 reads), allele frequency (AF ≤ 5% in gnomAD), and predicted deleteriousness (CADD score ≥ 20). Variants that passed the filter were manually curated following the ACMG criteria. We retained VUS, likely pathogenic, and pathogenic variants.

#### Software and statistical analysis

All statistical analyses were performed using R version 4.5.2 (2025-10-31) [32]. Data manipulation was conducted using dplyr v1.1.4 [33] and data.table v1.17.8 [34]. Visualization was performed using ggplot2 v4.0.0 [35] with additional packages including patchwork v1.3.2 [36] for multi-panel figures, ggrepel v0.9.6 [37] for label positioning, wesanderson v0.3.7 [38] for color palettes, and ggh4x v0.3.1 [39] for specialized plot elements. Protein interaction networks were constructed and visualized using ggraph v2.2.2, igraph v2.1.4 and tidygraph v1.3.1 libraries. We used EncrichmentTable v2.0.7 for pathway enrichment visualization. Circlize R package v0.4.16 was used for creating circos plots [40]. Venn diagrams and upset plots were generated using VennDiagram v1.7.3 [41] and UpSetR v1.4.0 [42], respectively. Image processing used magick v2.9.0 [43]. Python v3.12.3 was used for data processing and matrix construction, with pandas v2.1.4 for data frame operations.

Statistical significance was set at P ≤ 0.05 for all analyses unless otherwise specified. Multiple testing correction was applied where appropriate using the Benjamini-Hochberg false discovery rate method.

Artificial intelligence tools, including Claude (Anthropic) and GitHub Copilot, were used for sentence refinement and script optimization.

## RESULTS

### Population structure reveals distinct ancestry profiles

Principal component analysis (PCA) of low-frequency and rare variant genotypes revealed clear population structure between the two cohorts **(Figure 2, Supplementary Figure 2)**. MTL participants formed a relatively homogeneous cluster (mean pairwise genetic distance = 0.076) (**Supplementary Table 2**), consistent with predominantly European ancestry. In contrast, PTP participants displayed greater genetic variance (mean pairwise genetic distance = 0.127), with some individuals clustering near or overlapping with the MTL cohort while others were distinctly separated. This corresponds to a 67% higher within-group genetic diversity among PTP participants compared to MTL participants. Disease status (PD, CTRL) did not drive the primary clustering pattern, as both patients and controls were equally distributed within each cohort.

**Figure 2.**
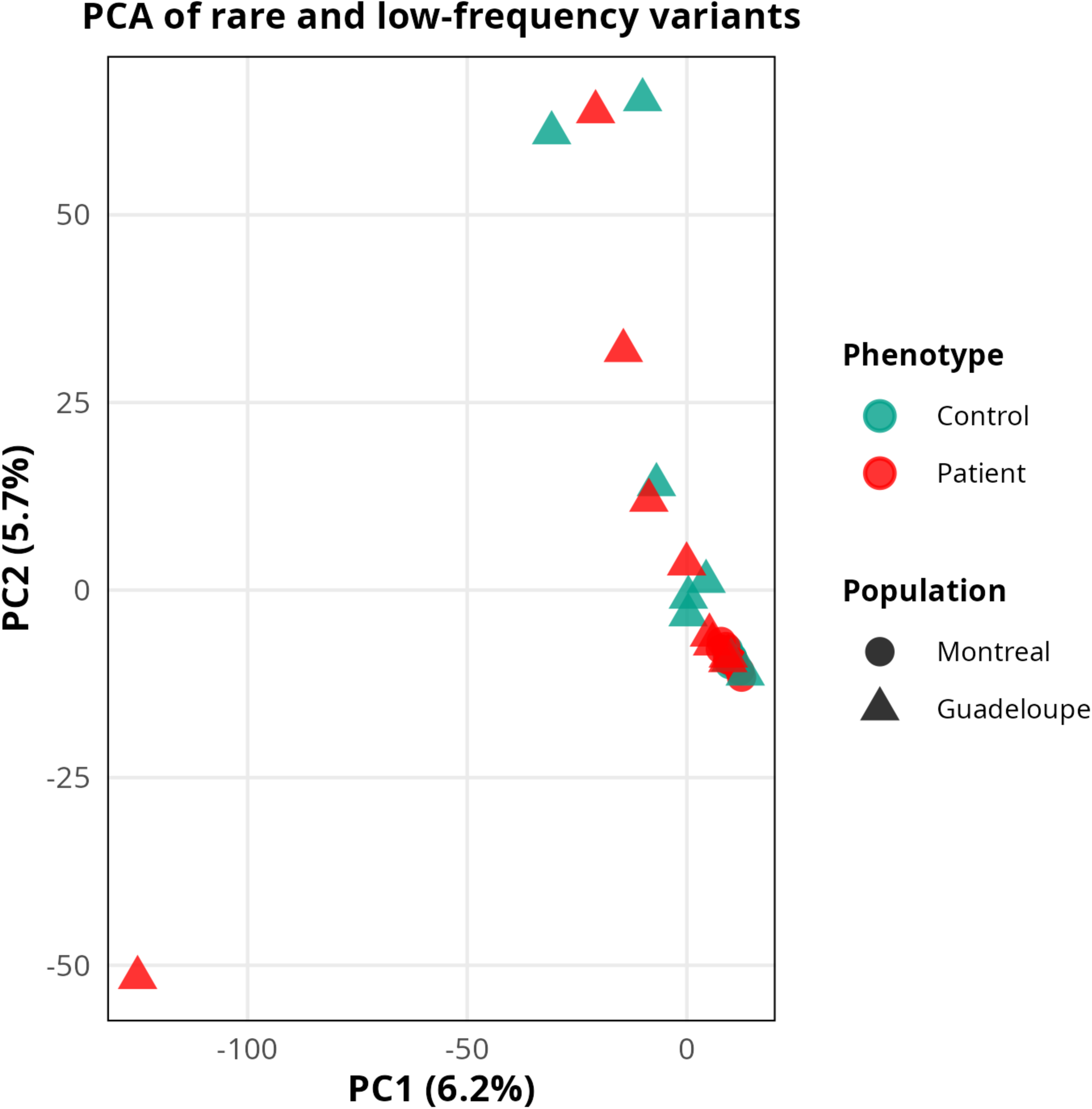
Principal component analysis reveals population structure in rare and low-frequency variants. PCA plot of rare (AF ≤ 0.05) and low-frequency (AF ≤ 0.1) variants demonstrates population structure across PC1 (6.2% variance) and PC2 (5.7% variance). Participants are grouped by study site: Montreal (circles) and Guadeloupe (triangles), with red indicating Parkinson’s disease patients and teal indicating controls.Montreal participants cluster tightly together, while Guadeloupe participants show greater dispersion across the PCA space, with some clustering with Montreal participants and others distributed more broadly. Cases and controls do not show systematic separation within the recruitment site.

Together, these findings confirm ancestry as the dominant source of genetic structure, thereby justifying ancestry-stratified burden analyses.

### Ancestry-stratified burden analysis reveals divergent immune mechanisms

#### IL-17 signaling pathway enrichment in Montreal case-control comparison

SKAT-C analysis identified genes with differential rare and low-frequency variant burden between our study groups. In the MTL cohort, 34 genes reached significance (P ≤ 0.05) **(Supplementary Table 3)**. The top hits included *RREB1* (P = 0.0047), a transcription factor involved in Ras signaling [44]; *MADD* (P = 0.005), which activates the mitogen-activated protein kinase ERK2 [45]; *SYNE1* (P = 0.006), which encodes a nuclear envelope protein that links the nucleus to the actin cytoskeleton [46]; *MIDEAS* (P = 0.01), involved in transcriptional regulation [47]. **(Figure 3A)**

**Figure 3.**
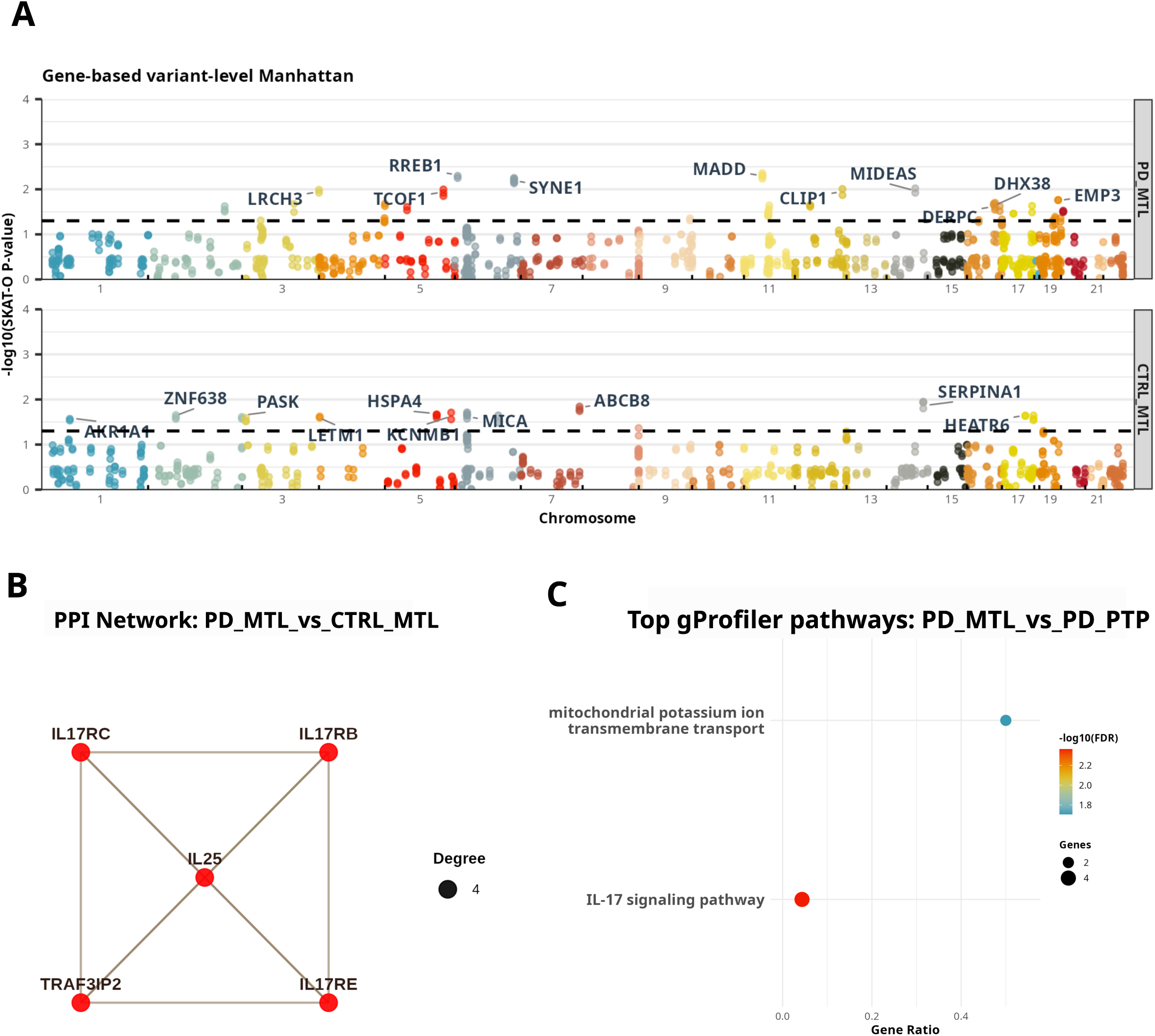
Ancestry-specific analysis in Montreal participants implicates IL-17 inflammatory signaling in Parkinson’s disease. **(A)** Gene-based Manhattan plot from SKAT-C burden testing comparing MTL_PD vs MTL_CTRL (PD_MTL vs CTRL_MTL). The dashed line indicates genome-wide significance threshold (p ≤ 0.05). Top hits are labeled. **(B)** Protein-protein interaction network of significant genes reveals a tightly connected IL-17 signaling module comprising IL17RC, IL17RB, IL25 (also known as IL17E), TRAF3IP2, and IL17RE. Node size represents degree: larger nodes indicate higher connectivity. **(C)** Top enriched pathways from g:Profiler analysis showing IL-17 signaling pathway as the most significant hit, along with mitochondrial potassium ion transmembrane transport. Dot size represents the number of genes in each pathway, and x-axis shows gene ratio (proportion of significant genes in pathway).

To investigate functional relationships among significant genes, we conducted a PPI network analysis using STRING. Of the 35 significant genes, 11 formed a connected subnetwork under confidence thresholds. The resulting network included 16 genes (11 query and 5 added interactors) and revealed one main functional module centered on IL-17 signaling components. The query genes *IL17RC* and *TRAF3IP2* formed the core of this network, with STRING adding three additional IL17 pathway members (IL17RE, IL17RB, IL25) as key interactors, highlighting the functional coherence of this module. **(Figure 3B)**

Pathway enrichment analysis of the 34 significant genes further supported enrichment of immune-related pathways in the MTL comparison. Using the g:Profiler enrichment tool, KEGG pathway analysis identified significant enrichment in the IL-17 signaling pathway (FDR=0.04), involving *TRAF3IP2, AKR1A1* and *GPCPD1* [48]. GO analysis revealed two additional enriched terms: mitochondrial potassium ion transmembrane transport (GO biological process, FDR=0.01) including *ABCB8,* and *LETM1.* It also highlighted IL17 family cytokine binding and signal transduction across the membrane (GO molecular function, FDR=0.03) including *AKR1A1* and *GPCPD1*. STRING enrichment analysis identified SEFIR domain and IL-17 signaling as significant pathways in this comparison. **(Figure 3C, Supplementary Table 4)**

The convergence of multiple levels of analysis implicates IL-17 signaling, increasing confidence that this represents a true biological signal rather than gene-level variation.

#### MHC class II pathway enrichment in Guadeloupe case-control comparison

In the PTP cohort, 40 genes reached significance **(Supplementary Table 3)**. The top hits included *RARG* (P = 0.007), a transcriptional regulator involved in retinoic acid signaling [49]; *HLA-DRB1* and *HLA-DQA1* (P = 0.01 for both), which encode major histocompatibility complex (MHC) class II molecules expressed on antigen-presenting cells [50]; *BST1* (P = 0.034), which promotes pre-B lymphocyte growth and has been implicated in rheumatoid arthritis pathogenesis [51,52,53] **(Figure 4A).**

**Figure 4.**
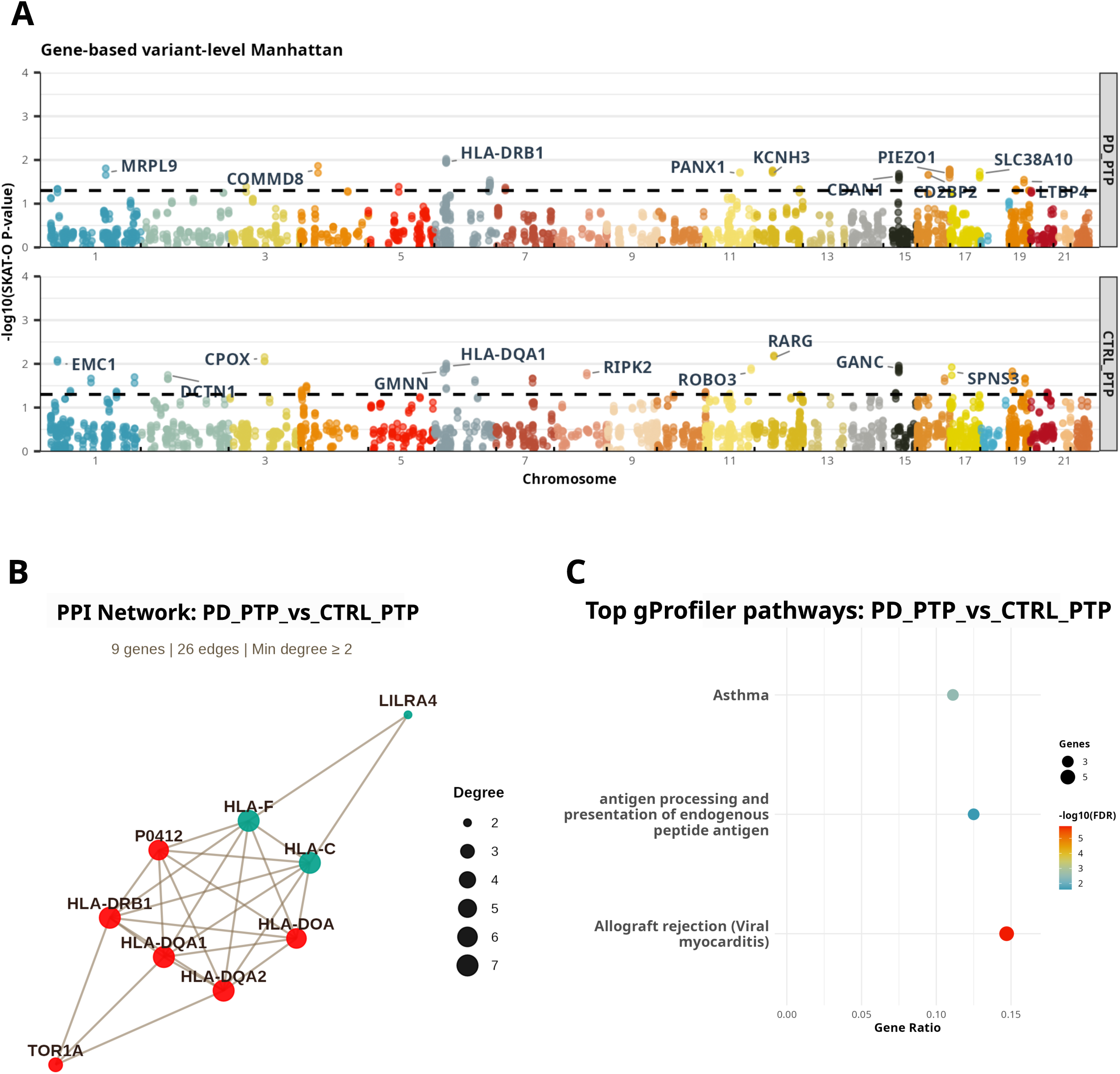
MHC-mediated adaptive immune mechanisms distinguish Guadeloupe Parkinson’s disease patients. **(A)** Gene-based Manhattan plot from SKAT-C burden testing comparing PD_PTP vs CTRL_PTP. Dashed line indicates significance threshold (p ≤ 0.05). Multiple HLA loci show strong association signals. **(B)** Protein-protein interaction network of significant genes is dominated by MHC class I (*HLA-F, HLA-C*) and class II molecules (*HLA-DRB1, HLA-DQA1, HLA-DQA2*), with additional connections to TOR1A, P0412, and LILRA4. Node size represents degree: larger nodes indicate higher connectivity. **(C)** Pathway enrichment analysis reveals immune-related processes including antigen processing and presentation of endogenous peptide antigen, allograft rejection, viral myocarditis, and asthma.

The PPI network included 23 genes (18 query + 5 STRING-added), with a densely connected HLA-centered module. Three HLA genes from SKAT-C (*HLA-DRB1, HLA-DQA1, HLA-DQA2*) formed the core, with STRING adding three additional MHC components (*HLA-C, HLA-F, HLA-DOA*) to complete the antigen presentation function **(Figure 4B).**

GO cellular component pathway enrichment revealed a significant signal of MHC protein complex (FDR = 1.59e-07), including *HLA-DQA2, XRCC1, MOGS, CHI3L2,* and *BST1*. Particularly, Wikipathways and Kyoto Encyclopedia of Genes and Genomes (KEGG) merged pathway terms, encompassed not only allograft rejection, interferon-ɣ (IFN-ɣ) signaling and autoimmune disease pathways, but also viral infection responses, viral myocarditis, and Human T-cell leukemia virus 1 infection. Asthma pathway was also significant in KEGG network while GO molecular function reported enrichment in MHC class II receptor activity **(Figure 4C, Supplementary Table 4)**.

These analyses consistently pointed toward MHC class II and IFN-related pathways, supporting coherent biological mechanisms rather than statistical noise.

#### Cross-ancestry PD comparison implicates divergent RNA processing and nuclear envelope architecture

The cross-ancestry comparison identified 89 genes with differential burden **(Supplementary Table 3)**. The top hits included *CD2BP2* (P = 0.0008), involved in pre-mRNA splicing and regulation of T lymphocyte activation [54], *MIDEAS* (P = 0.0018), involved in transcriptional regulation [47], *KCNH3* (P = 0.0029) which encodes a voltage-gated potassium channel [55], and *NUMA1* (P = 0.0029), involved in mitotic spindle organization [56] **(Figure 5A)**.

**Figure 5.**
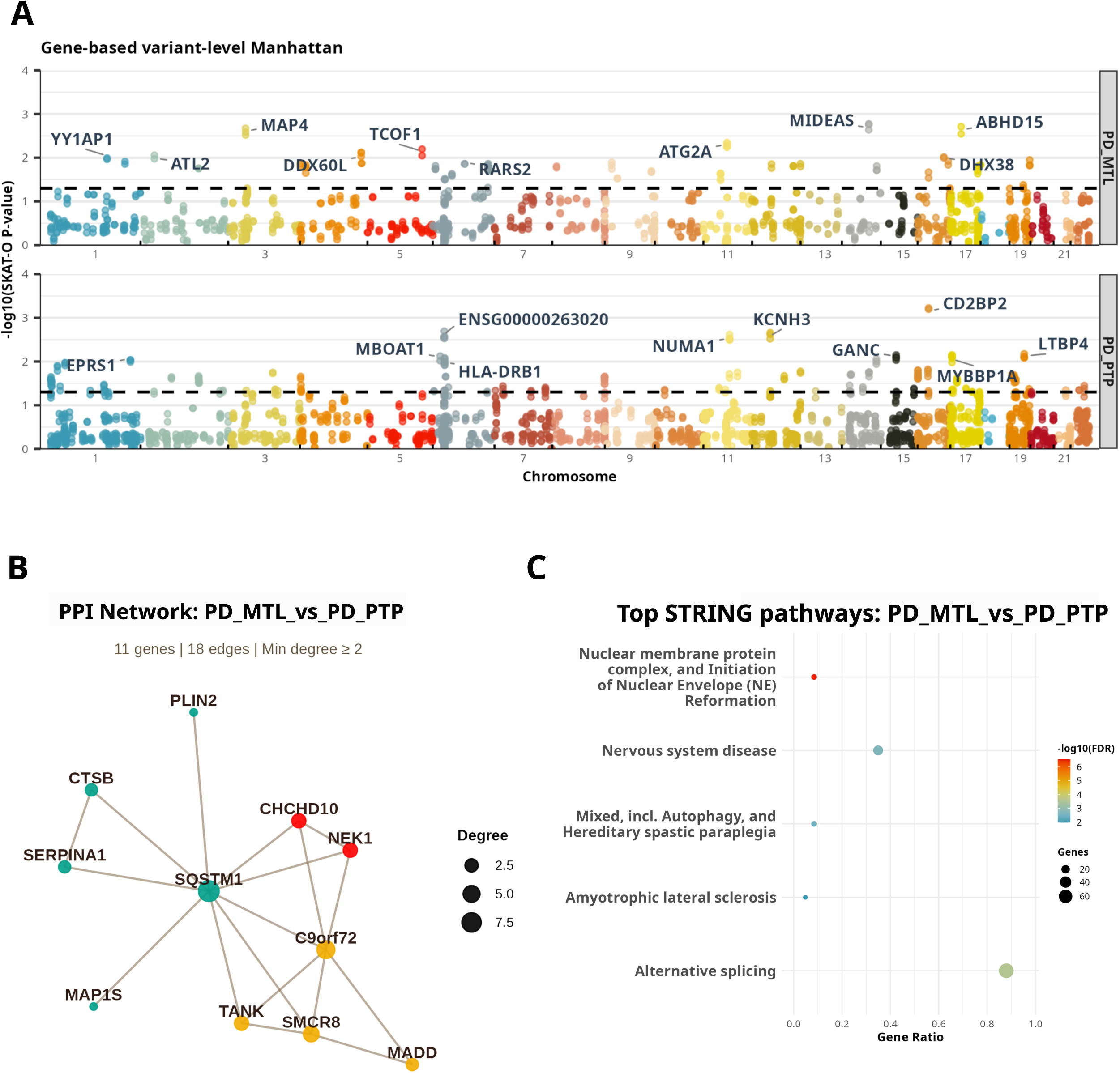
Direct comparison between ancestries reveals alternative splicing and nuclear envelope differences. **(A)** Gene-based Manhattan plot from SKAT-C burden testing directly comparing PD_MTL vs PD_PTP. Significant hits include genes involved in splicing regulation and nuclear membrane function. **(B)** Protein-protein interaction network of differentially enriched genes shows connections between splicing factors (SQSTM1, TANK, SMCR8, MADD) and nuclear envelope proteins (NEK1, CHCHD10, C9orf72), along with SERPINA1, CTSB, PLIN2, and MAP1S. **(C)** STRING pathway enrichment reveals alternative splicing as the predominant functional category, along with nuclear membrane protein complex/nuclear envelope reformation, nervous system disease, and pathways related to autophagy and hereditary spastic paraplegia. This suggests ancestry-specific differences in post-transcriptional regulation and neuronal homeostasis mechanisms.

The PPI network contained 49 genes (44 query + 5 STRING-added), with 11 highly connected proteins. STRING added *C9orf72* and *SQSTM1* as central hub genes connecting multiple SKAT-C significant genes. Both are established neurodegeneration genes: *C9orf72* harbors the most common genetic cause of familial amyotrophic lateral sclerosis/frontotemporal dementia [57,58], while *SQSTM1* encodes the autophagy adaptor protein p62, implicated in multiple neurodegenerative diseases [59,60] **(Figure 5B)**.

STRING enrichment analysis of the network identified multiple converging pathways. The network showed strong enrichment in alternative splicing (FDR = 2.6e-4), hematopoietic system (FDR = 1.3e-5), and nervous system disease (FDR=0.002). Nuclear envelope components demonstrated significant enrichment (FDR=2.9e-7) with protein including SYNE1, SYNE3, SYNE4, and PLEC. In accordance with the PPI analysis, the network revealed enrichment in autophagy and hereditary spastic paraplegia (FDR = 0.003), amyotrophic lateral sclerosis (FDR = 0.01), and neurodegeneration (FDR = 0.01) **(Figure 5C, Supplementary Table 4)**.

These differences suggest that distinct upstream immune processes may converge on shared neurodegenerative outcomes.

Canonical monogenic PD genes did not reach statistical significance in either ancestry group.

### Ancestry-specific haplotype structure mirrors population admixture in Guadeloupe

To investigate whether variants from SKAT-C-significant genes exhibited ancestry-specific LD patterns, we calculated LD (R²) between the SKAT-C variants from PD_ALL vs CTRL_ALL comparison and variants from 1000 Genomes Project Phase 3 reference panels, using both European (EUR) and combined African-admixed American (AFR-AMR) reference populations.

The analysis revealed substantial differences in LD architecture between ancestry groups. Among LD relationships identified, 23 variants pairs exhibited strong LD in both EUR and AFR-AMR reference panels, while 14 variant pairs showed AFR-AMR-specific LD patterns including those in *ATRIP, PIEZO1, ATP2A3,* and 11 variant pairs without being exhaustive in *RHBD1, RICTOR, NNT,* displayed EUR-specific LD patterns **(Figure 6A)**. Moreover, the 14 AFR-AMR-specific LD pairs were carried exclusively by PTP participants, with zero MTL participants harboring these LD relationships. In contrast, the 11 EUR-specific LD pairs were present in participants from both MTL and PTP cohorts, demonstrating genetic admixture in the PTP population. Importantly, PTP participants carrying AFR-AMR-specific LD pairs corresponded to those individuals who showed greater separation from the MTL cluster in principal component analysis **(Figure 2)**, while PTP participants carrying EUR-specific LD patterns were those who clustered closer to MTL participants **(Supplementary Table 5)**. This convergent evidence from LD analysis and population structure demonstrates that the PTP cohort encompasses a spectrum of African-European admixture proportions, with some individuals carrying predominantly African-ancestry haplotypes and others showing European genetic contributions.

**Figure 6.**
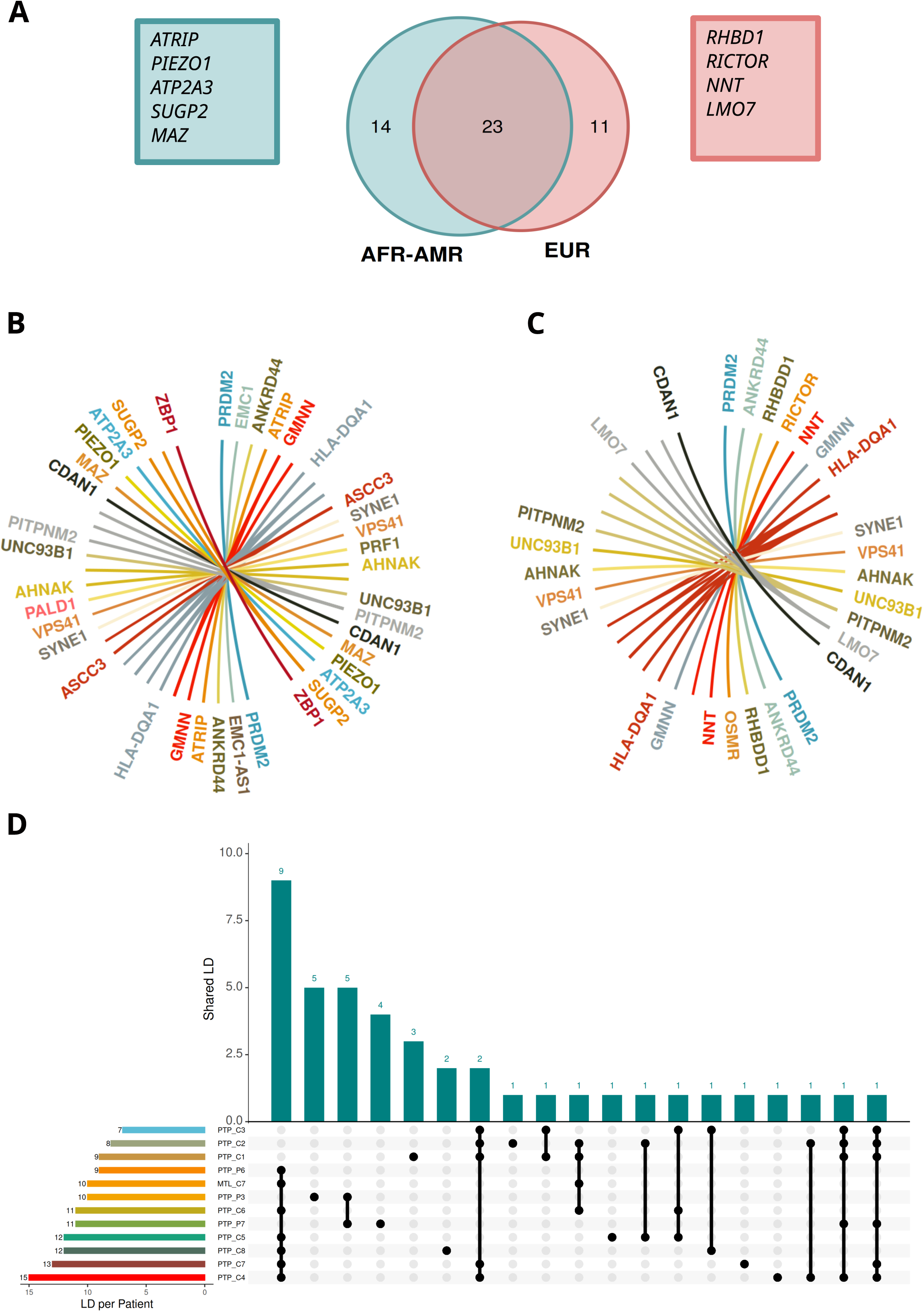
Linkage disequilibrium (LD) analysis reveals ancestry-specific and shared genetic architecture. **(A)** Venn diagram comparing significant genes identified in African-American /Admixed American (AFR-AMR) vs European (EUR) reference LD blocks from 1000 Genomes Project. Twenty-three genes show evidence of LD in both ancestry groups, while 14 are specific to AFR-AMR and 11 to EUR. Genes unique to each ancestry are listed in boxes. **(B-C)** Circos plots displaying LD relationships (R² > 0.2, 1000 kb window) between study variants (left semi-circle) and variants from the 1000 Genomes reference panel (right semi-circle), calculated using (B) AFR-AMR reference panel and (C) EUR reference panel. Each link represents an LD relationship between a single study variant and a single reference variant. Link thickness corresponds to LD strength (R² value). Links color indicates gene identity, with multiple links of the same color representing multiple variants from the same gene in LD. The distinct LD architectures observed between AFR-AMR and EUR reference analyses reflect ancestry-specific linkage patterns. **(D)** Left colored bars show individual LD burden per participant (height = number of LD pairs). Top bars indicate LD pairs shared between participants, with connecting dots/lines showing which participants share each LD pattern.

Circos plot visualization of LD networks revealed the bipartite structure of these relationships, with study variants (left semicircle) connected via links to 1000 Genomes reference variants (right semicircle). The distinct patterns observed between AFR-AMR **(Figure 6B)** and EUR **(Figure 6C)** reference panels reflect the different haplotype structures characteristic of each ancestral population. In total, our cohort exhibits 48 unique LD pairs, with 77.1% shared by multiple participants, while 22.9% were patient-specific. Top 10 carriers were predominantly PTP participants (7/10) and overall, PTP cohorts carried nearly three times more LD pairs than MTL participants. Three LD pairs were carried by ≥10 patients: 6:32641477-6:32641478 - 2 MTL and 12 PTP carriers (50% of cohort); 6:24781491-6:24781507 - 11 carriers all from PTP (39% of cohort); 11:62529063-11:62529129 - 4 MTL and 6 PTP carriers (36% of cohort). These ancestry-specific LD architectures further illustrate differences in haplotype organization between populations **(Figure 6D, Supplementary Table 5).**

We found no LD pairs between our variants and known GWAS PD risk variants, suggesting that the variants identified in this study represent independent genetic signals not captured by common variant association studies.

### Clinical phenotypes reflect ancestry-associated molecular divergence

Clinical assessments revealed differences in disease presentation between MTL and PTP participants. PD_MTL exhibited more advanced motor symptoms, with higher scores for axial rigidity, bradykinesia, and postural instability, along with greater overall disability as measured by the Hoehn & Yahr scale. PD_MTL also reported higher prevalence of prodromal features including REM sleep behavior disorder (71.4% vs 37.5%) and hyposmia (54.3% vs 22.5%). In contrast, PD_PTP displayed more autonomic symptoms, particularly orthostatic hypotension **(Supplementary Table 1)**. These clinical differences further support ancestry-associated heterogeneity in PD across both genetic and phenotypic levels.

### Exome sequencing identifies *NOD2* frameshift variant in Montreal PD patients

Our cohort comprised patients carrying variants of unknown significance (VUS) in known PD genes, including *PINK1* (NC_000001.11:g.20644665A>T), *PARK7* (NC_000001.11:g.7962772C>A), and *DNAJC6* (NC_000001.11:g.65385740G>A), each identified in one respective patient. Two patients carried likely pathogenic heterozygous *GBA* variants (NC_000001.11:g.155237458A>C, NC_000001.11:g.155238519T>G) [61].

Most variants in risk-factor and GWAS genes were classified as variants of uncertain significance (VUS) or likely benign/benign. The only pathogenic variant identified was a frameshift deletion in *CHRNB1* (NC_000017.11:g.7455868del) [11]. Three out of eight patients carried the same frameshift insertion in *NOD2* (NC_000016.10:g.50729870dup, p.Leu1007ProfsTer2), a gene previously reported as genome-wide significant in PD and a key innate immune receptor involved in NF-κB activation [11] (**Supplementary Table 6)**. In silico prediction tools indicated high likelihood of deleteriousness of this variant (CADD = 33), and it was classified as a VUS per ACMG/AMP criteria as implemented in Franklin (Genoox) [62].

### Control-only comparison confirms disease-specificity of enriched pathways

To distinguish disease-specific genetic associations from ancestry-related variation, we performed a control-only comparison (CTRL_MTL vs CTRL_PTP) **(Supplementary Table 3; Supplementary Figure 3A)**. This analysis identified 73 genes with significant differences in variant burden. Notably, this gene set included *NOD2* and *HLA-DQA1*, which also appeared in disease comparisons, indicating these genes harbor ancestry-related polymorphisms independent of disease status. However, these control-associated genes showed no evidence of functional coherence: PPI network analysis revealed only minimal connectivity (9 genes, 10 edges; **Supplementary Figure 3B**), and pathway enrichment analysis yielded no significant results.

The absence of coherent pathway enrichment in controls serves as an internal negative control, indicating that the molecular signals observed in PD are not driven by population stratification alone, but instead reflect ancestry-dependent disease-associated mechanisms.

## DISCUSSION

### Population structure and genetic admixture in Guadeloupe

The Guadeloupe islands were originally inhabited by indigenous Arawak, and later by Carib peoples, then French colonists established permanent settlement in 1635, importing enslaved Africans to work on plantations [63]. This history has resulted in substantial genetic admixture in the contemporary population. A genetic study of 198 individuals in Guadeloupe reported predominantly African maternal lineages (85%) followed by Eurasian and Amerindian lineages, with a marked sexual asymmetry (44% European paternal versus 7% European maternal ancestry) [64]. Despite our limited sample size, our cohort recapitulated this admixture pattern. PCA showed that while some PTP participants were genetically close to individuals of European ancestry, others were distinctly separated, corresponding to individuals carrying AFR-AMR-specific LD patterns.

### Divergent immune pathway enrichment implicates ancestry-dependent inflammatory mechanisms in PD

In the MTL cohort, rare and low-frequency variant burden, PPI and pathway enrichment analyses converged on IL-17-related inflammatory signaling. IL-17 is a family of pro-inflammatory cytokines primarily produced by Th17 cells and acting at the interface of innate and adaptive immunity [65]. IL-17 signaling interacts with major inflammatory pathways, including tumor necrosis factor (TNF) and nuclear factor kappa B (NF-κB), and amplifies immune responses in chronic inflammatory and autoimmune conditions [66,67]. Our findings align with prior studies implicating IL-17 in PD pathophysiology across multiple populations, including elevated IL-17 expression in brain tissue from an independent Chinese cohort [68], IL-17A-mediated microglial activation in rodent models [69], and a single-cell transcriptomic study reporting IL-17 pathway enrichment in activated monocytes from PD patients in MTL [17]. Together, these observations provide biological context for our results and support the role of IL-17-related immune dysregulation in PD pathogenesis.

In contrast, the PTP cohort was characterized by enrichment of MHC class II-mediated antigen presentation and IFNɣ-related pathways. IFNɣ is a central antiviral cytokine produced during viral infection that induces MHC class II expression across multiple cell types via STAT1 signaling, promoting antigen presentation and immune activation [70,71] [72,73,74]. Beyond CD4^+^ T cell activation, MHC class II molecules also play important roles in immune regulation, including maintenance of peripheral tolerance and modulation of autoimmune susceptibility [75].

The significance of MHC-II pathways in the PTP cohort may reflect ancestry-associated immune architecture shaped by environmental pressures. Pathogen diversity has been identified as a major driver of HLA allele selection across human populations [76]. In the Caribbean, long-standing exposure to endemic arboviruses such as dengue, Zika, and chikungunya elicits strong IFN-driven responses, and these infections can be associated with central nervous system symptoms, including parkinsonism [77,78,79]. Although causal links cannot be established in this study, this epidemiological context suggests that historical pathogen exposure may have contributed to shaping immune genetic architectures enriched for MHC class II and IFN-related pathways. Such ancestry-associated immune configuration may modulate inflammatory responses in PD and contribute to the distinct molecular and clinical features observed in the PTP cohort.

### Cross-ancestry burden differences converge on splicing dysregulation

Cross-ancestry burden analysis revealed enrichment for splicing-related gene architecture (73/89), including cytoskeletal proteins (SYNE family, PLEC), and the spliceosome component DHX38. Given the role of splicing dysregulation in several neurodegenerative disorders [80], burden differences in genes subject to complex alternative splicing may contribute to population-dependent disease mechanisms through altered isoform expression or splicing regulation.

### Emerging evidence supports ancestry-dependent mechanisms across neurodegenerative diseases

Our findings align with emerging evidence that disease mechanisms vary by ancestry in neurodegenerative diseases. Recent ancestry-stratified analyses of postmortem brain tissue revealed that genetic ancestry primarily influences immune cells and vascular endothelial cells, with African ancestry genes showing upregulation in immune pathways linked to Alzheimer’s disease and stroke risk, while European ancestry genes demonstrated upregulation in PD-associated pathways [81]. Multi-ancestry PD genome-wide studies reveal that immune and mitochondrial pathways emerge as significant only when diverse populations are included [82], and African-specific *GBA1* variants absent in European populations have been identified [83]. Our results of differential immune pathway enrichment extend these observations to the functional pathway level, suggesting that ancestry-inclusive research is essential for understanding PD heterogeneity and developing equitable precision medicine approaches.

### Admixture-driven LD heterogeneity shapes variant tagging in the Guadeloupe cohort

Our LD analysis demonstrates that differences in LD architecture across ancestries are a defining feature of the PTP cohort and closely mirror underlying population structure. Rather than presenting a uniform genetic background, the PTP cohort spans a continuum of African-European admixture, resulting in heterogeneous haplotype configurations that are not adequately captured by European reference panels alone. The increased LD burden observed in the PTP cohort therefore reflects haplotype structure shaped by admixture, rather than a genome-wide increase in LD.

These ancestry-specific LD architectures have important implications for genetic association analyses, as they directly influence which variants tag functional alleles and how disease-associated signals are detected. In admixed populations, reliance on European reference panels risks missing or misrepresenting biologically relevant variation, particularly in immune-related pathways. Our results underscore the necessity of ancestry-matched reference panels for accurate interpretation of genetic burden and highlight LD structure as a key mediator of ancestry-dependent molecular signals in PD.

### RBD and autonomic symptom profiles align with ancestry-specific immune pathway enrichments

The clinical differences observed between cohorts may reflect, in part, divergence in immune-related molecular pathways identified at the genetic level. Immune signaling and sleep regulation are tightly interconnected, with experimental evidence showing a bidirectional relationship between inflammatory cytokines and REM sleep. In a rat model, REM sleep deprivation increased circulating IL-17 and other pro-inflammatory cytokines [84]. Elevated circulating IL-17 has also been reported in depressive patients with sleep disorders [85]. In this context, the higher prevalence of RBD observed in the PD_MTL cohort aligns with the enrichment of IL-17-related pathways identified in our genetic burden analyses, suggesting a potential role for IL-17-driven immune activation in sleep-related manifestations of PD.

Conversely, the predominance of autonomic symptoms in the PD_PTP cohort may be consistent with an immune architecture characterized by IFN-driven responses, as activation of adaptive immune pathways has been shown to modulate autonomic regulation [86]. However, the specific mechanisms linking MHC-II pathway enrichment to autonomic dysfunction in PD remain to be elucidated.

While hypothesis-generating, this framework provides a biologically plausible hypothesis linking ancestry-associated immune variation to the distinct clinical phenotypes observed across cohorts.

### *NOD2* frameshift variant suggests a mechanistic link to IL-17 dysregulation in Montreal patients

Whole-exome sequencing identified a *NOD2* frameshift variant (NC_000016.10:g.50729870dup, p.Leu1007ProfsTer2) in three of eight Montreal PD patients, providing a potential mechanistic link to the IL-17 pathway enrichment observed in the PD_MTL cohort. NOD2 is an intracellular receptor that recognizes muramyl dipeptide (MDP), a component of bacterial peptidoglycan, and activates downstream NF-κB signaling and modulation of IL-17 production [87].

The NC_000016.10:g.50729870dup frameshift truncates the leucine-rich repeat domain, which is critical for ligand recognition and signaling [88]. This variant is a well-established risk factor for Crohn’s disease, although the functional consequences of this variant in PD have not been previously reported [89]. None of these patients had a history of inflammatory bowel disease or other autoimmune conditions.

While this variant does not meet ACMG pathogenicity criteria due to population frequency (AF ∼ 2% in European populations) and the presence of homozygous carriers in population databases, it may still contribute to disease susceptibility in the context of complex polygenic disorders such as PD. Under our study framework, which retains variants with AF ≤ 5% to capture risk-modifying alleles, the *NOD2* frameshift may represent one such allele contributing to IL-17 pathway dysregulation in a subset of patients, reinforcing the relevance of immune-mediated mechanisms in disease susceptibility.

## STRENGTHS AND LIMITATIONS

The multi-ancestry design of this study directly addresses a recognized gap in PD genetics research, where non-European populations remain severely underrepresented. The use of different analytical genetics approaches strengthens confidence that the identified immune signatures reflect disease-associated mechanisms rather than population stratification artefacts.

Nevertheless, the small sample size is the primary limitation: results are hypothesis-generating and do not support clinically meaningful conclusions without replication in larger cohorts. Future studies should seek replication in independent ancestry-matched cohorts and incorporate longitudinal follow-up to assess whether the identified molecular signatures track with disease progression.

## CONCLUSION

This study demonstrates ancestry-specific immune architectures in PD, with IL-17 signaling being predominant in European-ancestry patients and MHC class II/IFN pathways characterizing African-Caribbean patients. Although our discovery cohort is modest, the multi-level convergence of genetic, pathway enrichment, and clinical evidence strengthens confidence in these results and provides a robust foundation for hypothesis-driven follow-up studies. Ancestry-stratified rare-variant analyses therefore reveal mechanisms diluted in pooled populations, challenging a single-mechanism model of PD and supporting a precision-medicine framework in which inflammatory pathways differ across populations.

Future research should prioritize replication in diverse ancestry-inclusive cohorts, functional characterization of implicated variants in cellular and animal models, and longitudinal investigations of environmental exposures in relation to disease onset and progression.

Overall, our findings challenge a one-size-fits-all paradigm in PD genetics. The identification of distinct immune pathways underlying clinically overlapping phenotypes supports the emerging view that PD represents a spectrum of molecularly distinct disorders rather than a single pathogenic entity, with shared clinical manifestations arising from partially convergent biological pathways. Embracing this complexity through systematic investigation of underrepresented populations is essential for advancing mechanistic understanding and for achieving equity and efficacy in precision medicine for neurodegenerative disorders.

## Data Availability

All data produced in the present study are available upon reasonable request to the authors

## DECLARATIONS

### Ethics approval and consent to participate

The ethical boards of the Centre Hospitalier de l’Université de Montréal (CHUM) (2021-9115, 20.367-YP) and the University Hospital of Guadeloupe (Sud-Est I, 2020-88) approved the study. All participants provided written informed consent.

### Availability of data and materials

RNA-sequencing and exome-sequencing data have been deposited in the Gene Expression Omnibus (GEO) under accession number GSE325459 and the Sequence Read Archive (SRA) under BioProject accession PRJNA1439818, respectively. Custom scripts for bioinformatics analyses are available at https://github.com/TetreaultLab/ShortReadSequencing and https://github.com/TetreaultLab/Parkinson_variant_burden_analysis.

### Authors’ contributions

L.A designed the study, recruited participants, performed wet-lab processing, conducted bioinformatics analyses, and wrote the manuscript. M.L conducted bioinformatics analyses and revised the manuscript. L.Y.A.I performed variant curation and revised the manuscript. B.T performed statistical analysis of the clinical data and revised the manuscript. J.C recruited participants and revised the manuscript.

H.C performed wet-lab processing and revised the manuscript. S.G recruited participants and coordinated sample shipping from Guadeloupe and revised the manuscript. S.R has processed the PBMC samples in Guadeloupe and revised the manuscript. A.V.C performed clinical data analysis and revised the manuscript. S.C recruited participants, performed clinical assessments, and revised the manuscript. A.D recruited participants, performed clinical assessments, and revised the manuscript. MP contributed to study design, recruited participants, performed clinical assessments, contributed to grant application, and revised the manuscript. A.L contributed to study design, recruited participants, performed clinical assessments, contributed to grant application, and revised the manuscript. M.T contributed to study design, contributed to grant application, and supervised the research and revised the manuscript. All authors read and approved the final manuscript.

## Acknowledgements

We sincerely thank all the patients who participated in this study. We also thank UTMAB and University Hospital of Guadeloupe for their contribution to patients’ recruitment. Finally, we thank the sequencing facility at CHU de Quebec - Université Laval for sequencing services and the Digital Research Alliance of Canada for computational resources.

