## Supplemental figure 1 for "Ancestry-specific immune signatures in Parkinson’s disease: a rare variant burden analysis in Montreal and Guadeloupe cohorts"

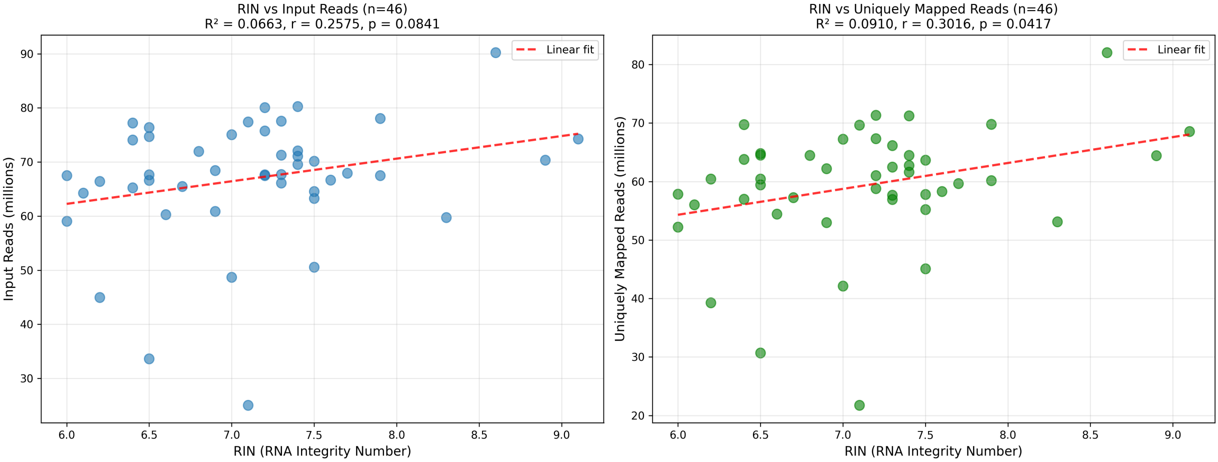


**Supplementary figure 1.** Correlation between RNA integrity number and sequencing depth across Montreal and Guadeloupe cohort.
