## Supplemental figure 2 for "Ancestry-specific immune signatures in Parkinson’s disease: a rare variant burden analysis in Montreal and Guadeloupe cohorts"

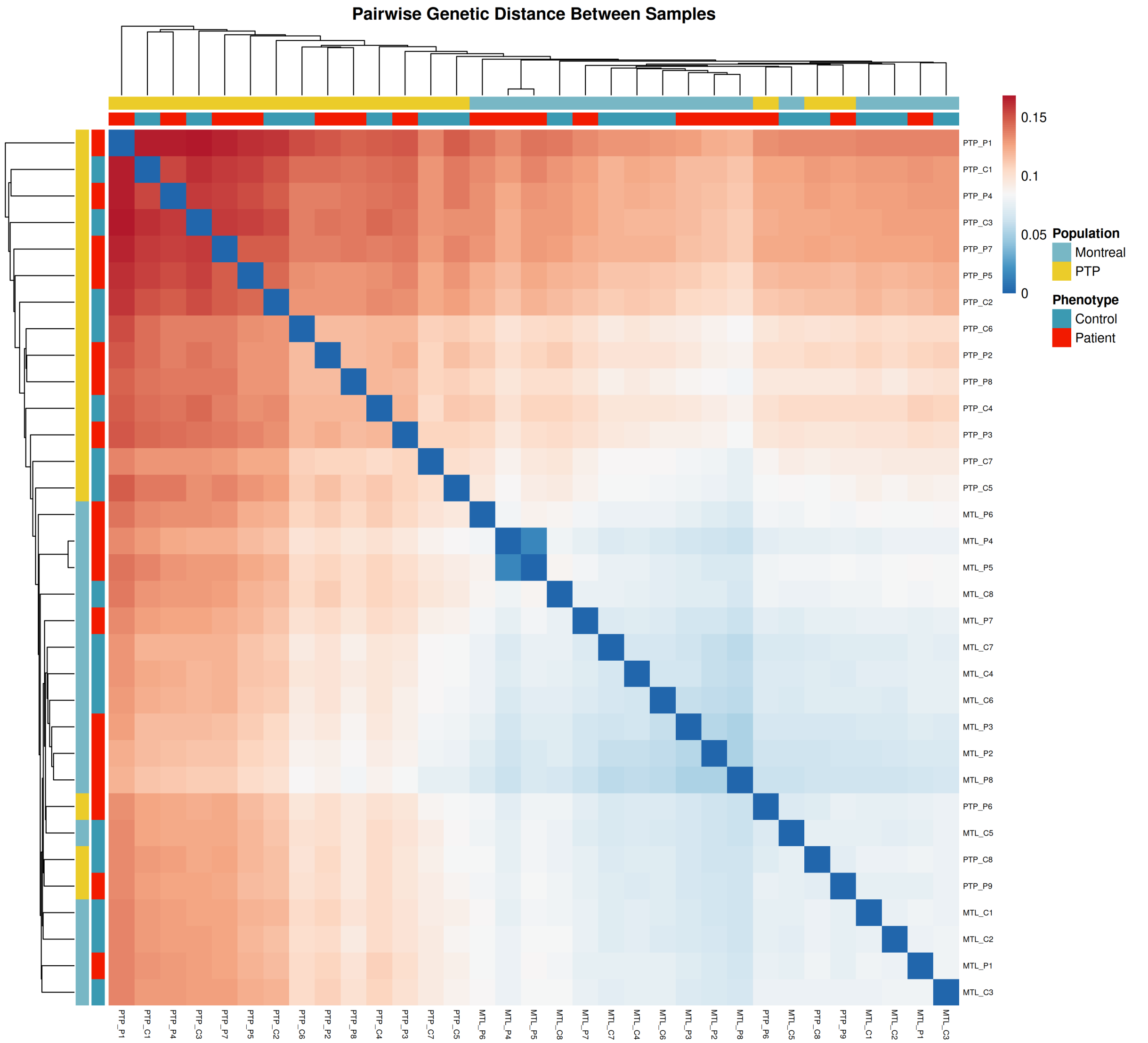


**Supplementary figure 3**. Hierarchical clustering of genetic distances between samples. Montreal participants cluster homogeneously while Guadeloupe participants show greater diversity consistent with admixture. Disease status does not drive clustering.
