## Supplemental figure 3 for "Ancestry-specific immune signatures in Parkinson’s disease: a rare variant burden analysis in Montreal and Guadeloupe cohorts"

A

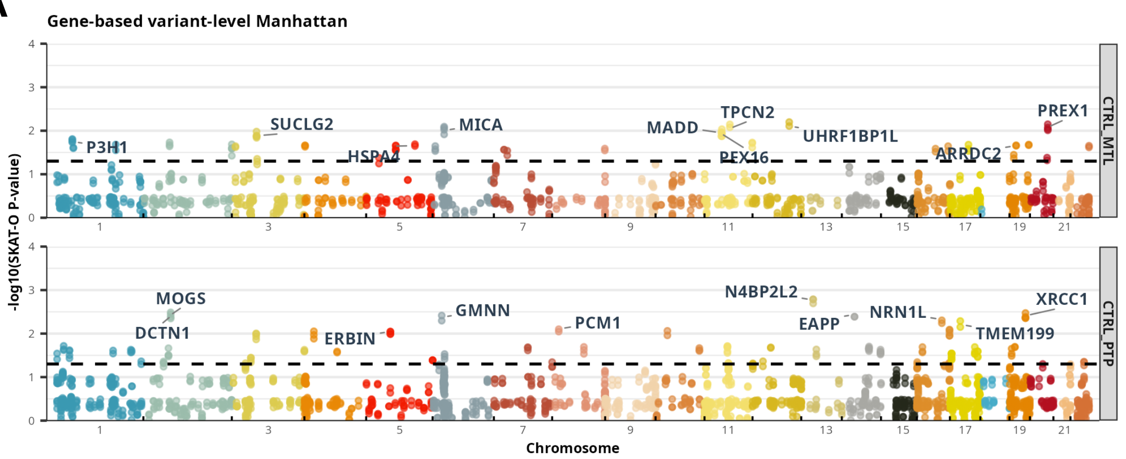


B

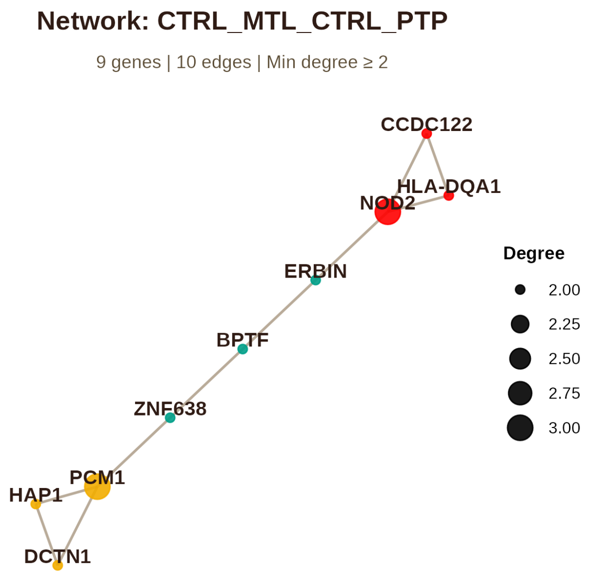


**Supplementary figure 2. (A)** Gene-based Manhattan plot from SKAT-C burden testing comparing CTRL_MTL vs CTRL_PTP. **(B)** Protein-protein interaction network of significant genes from SKAT-C results.
